# REM sleep EEG slowing signals basal forebrain integrity in older adults with subjective and mild cognitive impairment

**DOI:** 10.64898/2026.09.30.26363850

**Authors:** Aaron Lam, Nicole Espinosa, Elie Matar, Nathan Cross, Jurgen Fripp, Ying Xia, Claire André, Elizabeth Coulson, Renata Taranto, Ronald Grunstein, Angela L D’Rozario, Sharon L Naismith

## Abstract

The basal forebrain is an early site of pathology in Alzheimer’s disease (AD). Within this region, the nucleus basalis of Meynert (NbM) provides the primary cholinergic input to the neocortex and supports cortical activation during rapid eye movement (REM) sleep. REM electroencephalography (EEG) slowing is observed in older adults with AD and amnestic mild cognitive impairment (MCI). This study examined the relationship between REM EEG slowing and NbM volume, and whether it differed by cognitive status, sex or sleep apnea severity.

Participants aged ≥60 years with subjective cognitive impairment or MCI underwent neuropsychological and medical assessments, polysomnography and magnetic resonance imaging (MRI). REM EEG slowing was quantified as the ratio of delta and theta (<8Hz) to alpha, sigma and beta (8-32Hz) power. Bilateral NbM and Ch1-3 volume were derived from T1-weighted MRI images. Multiple linear regression and moderation analyses examined associations between REM EEG slowing and NbM volume, adjusting for age, sex, and sleep apnoea severity. To assess anatomical specificity, primary analyses were repeated using Ch1– 3 volume and non-REM EEG slowing. Exploratory analyses examined associations between NbM volume, sleep macroarchitecture, and memory.

The sample comprised 116 participants (59.5% female; mean age = 70.5 years, 64.7% MCI). Greater REM EEG slowing was associated with smaller NbM volume (all p < 0.05), whereas no significant associations were observed with Ch1–3 volume and non-REM EEG slowing (all p > 0.05). Although moderation effects were not significant after FDR correction, within-group analyses suggested a stronger relationship in MCI. Smaller NbM volume was associated with longer sleep latency and poorer episodic memory (all p < 0.05).

REM EEG slowing may reflect early NbM degeneration and provide a non-invasive biomarker of basal forebrain integrity in older adults with cognitive concerns, with findings showing anatomical specificity to the NbM and sleep-stage specificity to REM sleep. Longitudinal studies are required to establish its prognostic value.

## Introduction

The most common form of dementia is Alzheimer’s Disease, which is characterised by the pathological accumulation of amyloid-beta plaques and tau neurofibrillary tangles in the brain, leading to progressive neurodegeneration.^1^ These pathologies typically develop 10-20 years before the onset of clinical symptoms.^2^ The cholinergic system, particularly the basal forebrain (BF), is among the earliest neural systems affected in Alzheimer’s disease.^3^ The BF nuclei play a crucial role in memory and arousal.^4^ Within the BF, the nucleus basalis of Meynert (NbM) provides widespread cholinergic innervation to the neocortex and is particularly vulnerable to early Alzheimer’s disease-related neurodegeneration. Degeneration of NbM cholinergic neurons has been proposed as a potential early biomarker and therapeutic target for dementia.^5^

During rapid eye movement (REM) sleep, the NbM is highly active and contributes to cortical activation and electroencephalographic (EEG) desynchronisation, a distinctive feature of REM sleep compared to other sleep stages.^6^ Degeneration of NbM cholinergic neurons in Alzheimer’s disease may therefore contribute to alterations in REM sleep physiology. Individuals with Alzheimer’s disease typically exhibit reduced REM sleep duration and prolonged REM onset latency relative to cognitively intact older adults.^7^ Furthermore, characteristic changes in sleep microarchitecture, specifically pathological EEG slowing during REM sleep, have been documented in people with Alzheimer’s disease compared to healthy controls.^8^ EEG slowing during REM sleep refers to a shift in neural activity characterised by a reduction in fast frequencies, typically in the sigma (10-16Hz) and beta (16-32Hz) bands. This occurs alongside a concurrent increase in slow frequencies, particularly in the delta (0.5-4Hz) and theta (4-8Hz) bands. Importantly, these EEG changes have also been observed in ‘high risk’ or prodromal mild cognitive impairment (MCI) stages,^9^ supporting the notion that NbM-related cholinergic dysfunction during REM sleep may emerge early in the disease process.

Despite a strong physiological basis linking REM EEG slowing to cholinergic function, direct *in vivo* evidence remains limited. One study of 24 older adults with and without MCI^10^ used [¹ F]fluoroethoxybenzovesamicol (FEOBV) positron emission tomography (PET) to assess cholinergic innervation and found that REM EEG slowing was associated with widespread cholinergic denervation. Notably, comparable associations were not observed for EEG slowing during non-REM sleep or wakefulness, suggesting that REM EEG slowing may be particularly sensitive to cholinergic dysfunction rather than reflecting non-specific EEG slowing. However, associations between basal forebrain volume and REM EEG slowing did not survive correction for multiple comparisons. This may reflect the study’s limited statistical power and predominantly cognitively unimpaired sample. Complementing these molecular imaging findings, a recent study in Lewy body disorders found that greater occipital REM EEG slowing was associated with altered functional connectivity of key cholinergic pathways, most consistently the pedunculopontine nucleus (PPN)–thalamic pathway and, to a lesser extent, NBM–cortical networks.^11^ Together, these findings suggest that REM EEG slowing may be a marker of cholinergic function, although evidence linking REM EEG slowing to the structural integrity of the NbM remains limited. Furthermore, exploratory sex-stratified analyses suggested that the relationship between REM EEG slowing and cholinergic denervation may differ between women and men, pointing to possible sex differences in its neural basis.^10^ However, these analyses were exploratory, limited by small sex-stratified samples, and did not formally test a sex-by-REM-slowing interaction. Whether sex moderates the association between REM EEG slowing and NbM structural integrity therefore remains unknown.

Prior studies examining REM EEG slowing and cholinergic markers in older adults with MCI and dementia with Lewy Bodies tend to exclude obstructive sleep apnea (OSA) which limited the generalisability of existing findings to clinical memory-clinic populations.^10,11^ Our previous work indicates that up to 44% of older adults attending memory clinics have at least moderate OSA.^12^ OSA has been previously linked to changes in sleep EEG activity, including REM EEG slowing.^13,14^ Therefore, the extent to which OSA modifies or obscures the association between REM EEG slowing and NbM volume also remains unknown. Establishing whether this relationship persists is important for determining the clinical utility of REM EEG slowing as a marker of cholinergic system integrity.

Given the widespread cortical cholionergic projections of the NbM, its structural integrity also has functional significance for learning and memory processes. Prior work has shown that reduced NbM volume is associated with poorer cognitive performance for those with both MCI and Alzheimer’s disease, although findings regarding the contribution of NbM integrity to memory performance have been inconsistent, particularly at earlier stages of cognitive impairment [i.e. subjective cognitive impairment (SCI)].^15,16^ We therefore investigated whether lower NbM volume was associated with poorer learning and memory performance in older adults with SCI and MCI.

The present study aimed to investigate the relationship between quantitative EEG (qEEG) spectral features of REM sleep and NbM volume in older adults with SCI and MCI attending a brain health and memory clinic. First, we examined whether the REM EEG slowing ratio was associated with NbM volume in the whole sample. Second, we explored whether cognitive status, sex, or OSA severity moderated this relationship. Third, given the proposed role of the NbM in cortical cholinergic modulation during REM sleep, we also examined the anatomical and sleep-stage specificity of this association by testing Ch1–3 volume and non-REM EEG slowing as comparison measures. Finally, as secondary exploratory analyses, we examined whether NbM volume is associated with broader aspects of sleep macroarchitecture and learning and memory impairments.

We hypothesised that greater REM EEG slowing would be linked to lower NbM volume. We expected that cognitive status would moderate this relationship, such that the association between REM EEG slowing and NbM volume would be stronger in MCI, given more pronounced changes in both REM EEG and NbM volume at this clinical stage. Based on preliminary evidence that the anatomical correlates of REM EEG slowing may differ between women and men,^10^ sex was examined as a potential moderator without a prespecified directional hypothesis. We hypothesised that the primary association would demonstrate sleep-stage and anatomical specificity, such that the non-REM EEG slowing ratio would not predict NbM volume and the REM EEG slowing ratio would not predict Ch1-3 volume. Lastly, given the known effects of OSA on REM sleep architecture, we hypothesised that the association between REM EEG slowing and NbM volume would be attenuated with increasing OSA severity. Associations between NbM volume, sleep macroarchitecture and learning and memory performance were considered secondary and exploratory.

## Materials and methods

### Participants

As previously described,^17^ participants were recruited from the Healthy Brain Ageing (HBA) Clinic at the University of Sydney, Australia. The HBA Clinic is a specialised clinic for older adults concerned about recent changes to their cognition. Eligibility required participants to be aged 50 years or older, to be referred from either a general practitioner or a medical specialist and to have a Mini-Mental State Examination (MMSE)^18^ scores ≥20. Exclusion criteria comprised a previous diagnosis of dementia, psychiatric or neurological disorders (e.g. bipolar disorder, Parkinson’s Disease), intellectual disability, history of stroke or significant head trauma (loss of consciousness greater than 30 minutes), significant prior or current (≥14 standard alcoholic drinks per week) substance misuse, or insufficient English proficiency to complete the neuropsychology assessment. For the present study, we also exclude participants aged 50–59 years, those with mild dementia (MMSE < 24) and those whose sleep and neuroimaging assessments were conducted >180 days apart. Participants taking antidepressants or cholinesterase inhibitors were also excluded due to their documented effects on REM sleep physiology.^19,20^

Ethical approval was granted by the University of Sydney Human Ethics Committee (No. 2012/HE001873 and 2019/HE000271), and all participants provided written informed consent before any study procedures.

### Clinical and Memory Assessments

As previously detailed,^17^ participants underwent comprehensive clinical assessments, neuropsychological testing and mental health and sleep evaluations with a medical specialist, neuropsychologist and research psychologist respectively. Medical specialists collected detailed medical histories, medication use, body mass index (BMI; kg/m²) and weekly alcohol intake (standard drinks per week).

Through self-report measures, depressive symptoms were measured using the Geriatric Depression Scale short form-15 (GDS-15),^21^ where higher scores indicated more depressive symptoms (scores ranged from 0 to 15). Sleep quality was measured using the Pittsburgh Sleep Quality Index (PSQI)^22^, whereby higher scores indicating poorer sleep quality (scores ranged from 0 to 21).

Although a comprehensive neuropsychological test battery was administered,^17^ the primary cognitive outcomes examined in this study were verbal learning and memory. As described previously,^23^ this was assessed using two composite *z*-scores that were normalised for age and education. The *verbal learning composite* score comprised the five learning trials (trials 1-5) of the Rey Auditory Verbal Learning Test (RAVLT)^24^ and the summed encoding component of the Wechsler Memory Scale-III Logical Memory task.^25^ The *verbal memory composite* score comprised the delayed recall trial (trial 7) of the RAVLT and the Wechsler Memory Scale-III Logical Memory delayed recall task.

Via clinical consensus, participants were classified into cognitive impairment subtypes based on standardised criteria. MCI was defined according to the Winblad criteria, with neuropsychological test performance at least 1.5 standard deviations below age-specific normative data, relative to estimated premorbid IQ.^26^ Participants with MCI were further subtyped as amnestic MCI (aMCI) and non-amnestic (naMCI) according to predominant deficits in the memory *vs.* non-memory domains. Using Jessen criteria,^27^ SCI was defined as having subjective cognitive impairment but not meeting criteria for MCI.

### Neuroimaging and basal forebrain volume processing

Structural magnetic resonance imaging (MRI) data were acquired at two sites in Sydney, Australia. The first site used a 3-Tesla General Electric (GE) Discovery MR750 scanner (GE Medical Systems, Chicago, IL, USA). T1-weighted images were acquired using a 3D brain volume imaging (BRAVO) spoiled gradient-recalled sequence. Before July 2019 (*n* = 61), scans were obtained with an eight-channel phased-array head coil (TR = 7.2 ms, TE = 2.8 ms, flip angle = 12°, matrix = 256 × 256, 196 sagittal slices, 0.9 mm isotropic voxels, acquisition time = 4 min 27 s). From July 2019 onwards (*n* = 48), a 32-channel phased-array head coil was used with a slightly updated sequence (TR = 7.4 ms, TE = 3.0 ms, flip angle = 11°, matrix = 256 × 256, 1.0 mm isotropic voxels, acquisition time = 4 min 5 s). The second site (*n* = 19) used a 3T Siemens MAGNETOM Skyra scanner (Siemens Healthineers, Erlangen, Germany) equipped with a 20-channel phased-array head coil. T1-weighted images were obtained using a three-dimensional magnetisation-prepared rapid gradient-echo sequence, comprising 192 sagittal slices with the following acquisition parameters: repetition time = 2200 ms, echo time = 2.48 ms, flip angle = 8° and acquisition matrix = 248 × 256.

T1-weighted images were processed using the Computational Anatomy Toolbox (CAT12, version 12.8; www.neuro.uni-jena.de/cat/index.html) for SPM12 (https://www.fil.ion.ucl.ac.uk/spm) in MATLAB (R2018b; MathWorks, Natick, MA, USA). Images were corrected for intensity non-uniformity and segmented into grey matter, white matter and cerebrospinal fluid using the default CAT12/SPM12 pipeline. A study-specific DARTEL template was generated from all participants and grey-matter maps were spatially normalised to Montreal Neurological Institute (MNI) space using the corresponding deformation fields. Normalised grey-matter maps were resampled to 1 mm isotropic resolution without spatial smoothing. Total intracranial volume was estimated using CAT12.

Basal forebrain regions-of-interest (NbM and Ch1-3) were defined using probabilistic cytoarchitectonic masks^28^ and implemented using the JuBrain Anatomy toolbox.^29^ The masks were reliced onto the common 1-mm MNI grid using the nearest-neighbour interpolation. The bilateral NbM (Ch4) and Ch1-3 (medial septum, vertical and horizontal limbs of the diagonal band of Broca) masks were constructed from the union of the left and right masks. The bilateral NbM and Ch1-3 were normalised to total intracranial volume and expressed as the volume of the region of interest multiplied by 1 million, divided by the total intracranial volume. Visual quality control was performed on all raw T1 images, the CAT12 segmentation, the spatial normalisation and the region-of-interest overlays. Twelve participants were excluded due to poor image quality, segmentation failure, normalisation error, or misaligned region-of-interest placement, resulting in a final sample of N = 116 for analyses.

### Polysomnography and EEG analysis

Participants underwent a single-night polysomnography at the Woolcock Institute of Medical Research in Sydney, Australia. EEG signals were acquired from electrodes placed at standard locations (F3, Fz, F4, C3, Cz, C4, Pz, O1, O2) with contralateral mastoid referencing (midline referenced to the average of M1/M2). Data were sampled at frequencies of 128 Hz, 200 Hz, 256 Hz, or 512 Hz, depending on the system (Grael, Sandman Elite, Alice 5, REMLogic, respectively). Sleep stages and respiratory events were manually scored according to standard American Academy of Sleep Medicine (AASM) v2.3 criteria by trained sleep technicians.^30^ Participants were instructed to sleep between 9:00 and 10:30 PM, with an 8-hour sleep opportunity. OSA severity was assessed using the apnea-hypopnea index (AHI), with an AHI ≥ 15 events/hour indicating the presence of moderate or severe OSA and an AHI < 15 events/hour classified as no or mild OSA. For descriptive purposes, we report additional sleep macroarchitecture metrics including total sleep time, sleep efficiency, sleep onset latency, wake time after sleep onset, non-REM sleep duration, REM sleep duration, REM sleep onset latency, oxygen desaturation index (ODI) and the arousal index.

### Sleep microarchitecture measures

For each available EEG electrode, power spectral density was computed using a fast Fourier transformation for each consecutive 5-second epoch. The primary measure was the REM EEG slowing ratio. As previously described^9^, this was calculated by [delta (0.5–4.5 Hz) + theta (4.5-8 Hz)] / [alpha (8–12 Hz) + sigma (12–15 Hz) + beta (15–32 Hz)]. To ascertain the specificity of our associations, we also derived the same slowing ratio for non-REM sleep. Regional measures were derived for frontal (F3, F4, Fz), central (C3, C4, Cz), parietal (Pz) and occipital (O1 and O2) regions. Within each region, absolute power was first averaged across available electrodes for each frequency band, after which the regional slowing ratio was calculated. When one or more electrodes within a region were unavailable, the regional average was calculated from the remaining available electrodes.

### Statistical analysis

All statistical analyses were conducted using R version 4.4.2. Topographic plots were generated using the *ggplot2* package. Model assumptions were assessed by examining histograms and Q-Q plots of residuals for normality and homoscedasticity, Levene’s test for homogeneity of variance, scatterplots for linearity and variance inflation factors (VIF < 5 considered acceptable) for multicollinearity. Descriptive analyses examined differences in demographic and clinical characteristics, bilateral NbM and Ch1-3 volumes and sleep measures between cognitive status groups (SCI *vs.* MCI). Multiple linear regression models were used to examine whether REM EEG slowing at each electrode was associated with bilateral NbM volume. Unadjusted models were first fitted, followed by models adjusted for age, sex, OSA severity and MRI sequence. Interaction models then examined whether associations between REM EEG slowing and NbM volume were moderated by cognitive status, sex, or OSA severity, adjusting for age and MRI sequence. The Benjamini–Hochberg false discovery rate (FDR) procedure was applied to account for multiple comparisons across the four scalp regions. Statistical significance was defined as a two-tailed p < 0.05. To assess sleep-stage specificity, primary analyses were repeated using the non-REM EEG slowing ratio. To assess anatomical specificity, primary analyses were repeated using Ch1–3 volumes as outcomes.

Exploratory Pearson’s correlation analyses examined associations between NbM, Ch1-3, total basal forebrain volume and sleep macroarchitecture. In line with prior work^31^, we examined these relationships in the whole sample and in the aMCI subsample. Additional correlations examined associations between NbM volume and learning and memory performance. Spearman correlations were used where appropriate when data transformations did not achieve normality.

## Results

A total of 116 participants were included in the final analysis. The mean age was 70.5 years (with a range of 60 - 88 years) and 59.5% were female. Participants were generally highly educated (*M* = 13.6 years) and had a mean BMI within the ‘slightly overweight’ range. Clinically, 64.7% of participants met the criteria for MCI, including 25% with aMCI and 38.8% had moderate or severe OSA. Compared to participants with SCI, those with MCI consumed fewer standard units of alcohol (*p* < 0.05) and had smaller NbM and Ch1-3 volumes (both *p* < 0.05). Full demographic and clinical characteristics, including comparisons between the SCI and MCI groups, are presented in Table 1.

**Table 1.**
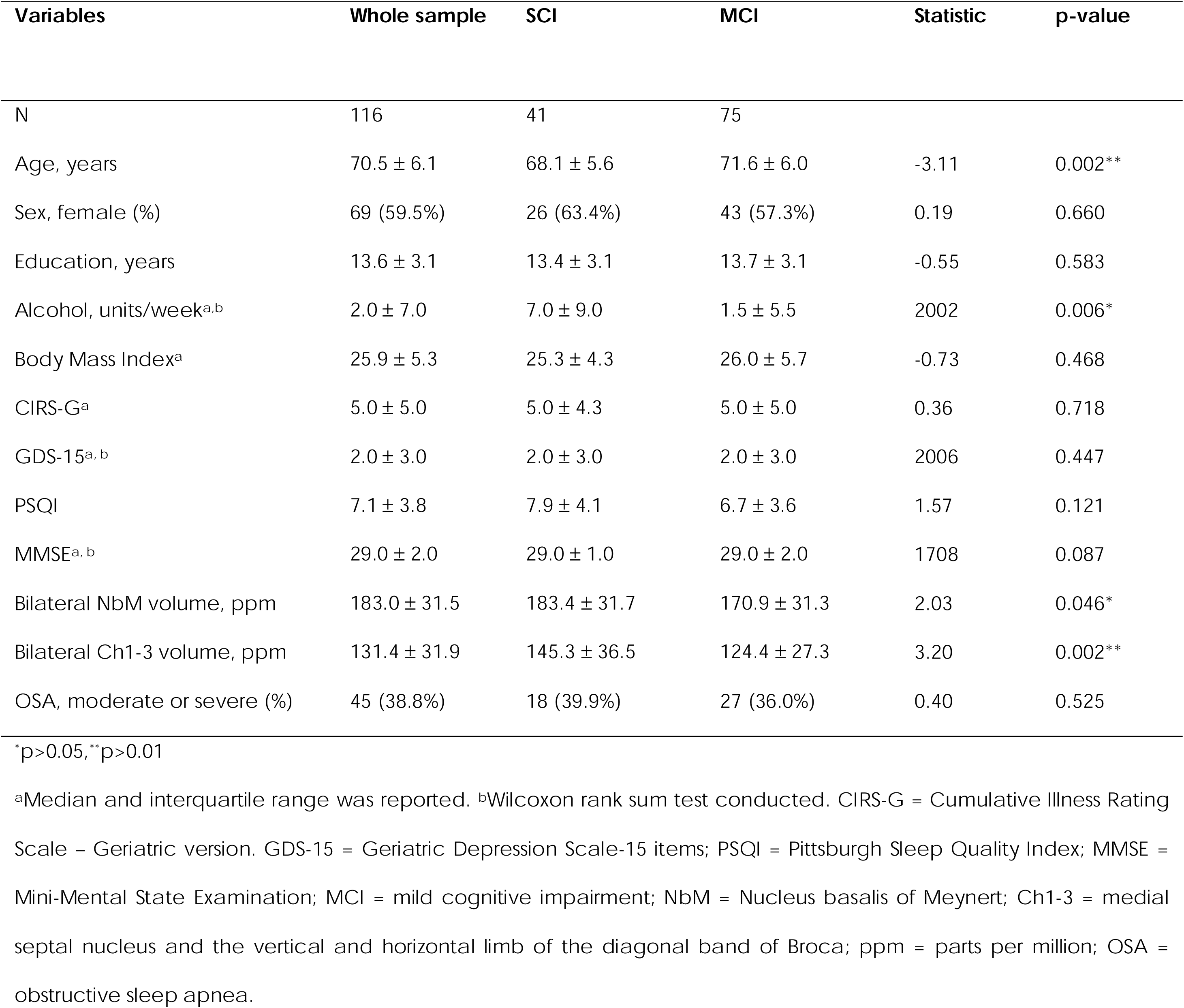
Participant demographics and clinical characteristics.

In terms of sleep macroarchitecture, there were no differences between SCI and MCI groups on any sleep duration and OSA severity measure (all *p* > 0.05; see Table 2). Across all regions, REM slowing ratio did not differ between the SCI and MCI groups (all *p* > 0.05; see Table 3).

**Table 2.** Sleep macroarchitecture profiles for the whole sample and clinical subjective cognitive decline (SCI) and Mild Cognitive Impairment (MCI) subgroups.

| Variables | Whole sample | SCI | MCI | Statistic | p-value |
| --- | --- | --- | --- | --- | --- |
| N | 116 | 41 | 75 |  |  |
| Total sleep time, min | 334.0 ± 73.2 | 334.0 ± 71.3 | 334.1 ± 71.9 | -0.01 | 0.994 |
| Sleep efficiency, %<sup>a,b</sup> | 73.2 ± 18.8 | 71.6 ± 15.2 | 74.3 ± 18.5 | 1416 | 0.485 |
| Sleep onset latency, min<sup>a</sup> | 15.5 ± 30.6 | 18.5 ± 41.8 | 14.0 ± 25.0 | 0.54 | 0.594 |
| WASO, min<sup>a</sup> | 75.3 ± 78.0 | 90.5 ± 88.0 | 74.5 ± 73.3 | 0.47 | 0.639 |
| Non-REM duration, % | 81.9 ± 7.0 | 80.6 ± 6.4 | 82.6 ± 7.2 | -1.55 | 0.125 |
| REM duration, % | 18.1 ± 7.0 | 19.4 ± 6.4 | 17.4 ± 7.2 | 1.55 | 0.125 |
| REM onset latency, min<sup>a</sup> | 86.8 ± 77.6 | 95.0 ± 73.9 | 82.5 ± 78.0 | 0.31 | 0.748 |
| AHI, n/hour<sup>a</sup> | 11.7 ± 14.8 | 9.6 ± 14.1 | 11.9 ± 14.8 | -0.68 | 0.496 |
| ODI, n/hour<sup>a</sup> | 5.2 ± 9.4 | 5.0 ± 12.7 | 5.4 ± 7.9 | -0.76 | 0.448 |
| Arousal index, n/hour<sup>a</sup> | 19.3 ± 15.2 | 20.1 ± 16.5 | 18.5 ± 13.0 | 0.29 | 0.774 |

\*p>0.05,\*\*p>0.01
<sup>a</sup>Median and interquartile range was reported. <sup>b</sup>Wilcoxon rank sum test conducted. OSA = obstructive sleep apnea; WASO = wake after sleep onset; REM = rapid eye movement; AHI = apnea hypopnea index; ODI = oxygen desaturation index.

**Table 3.**
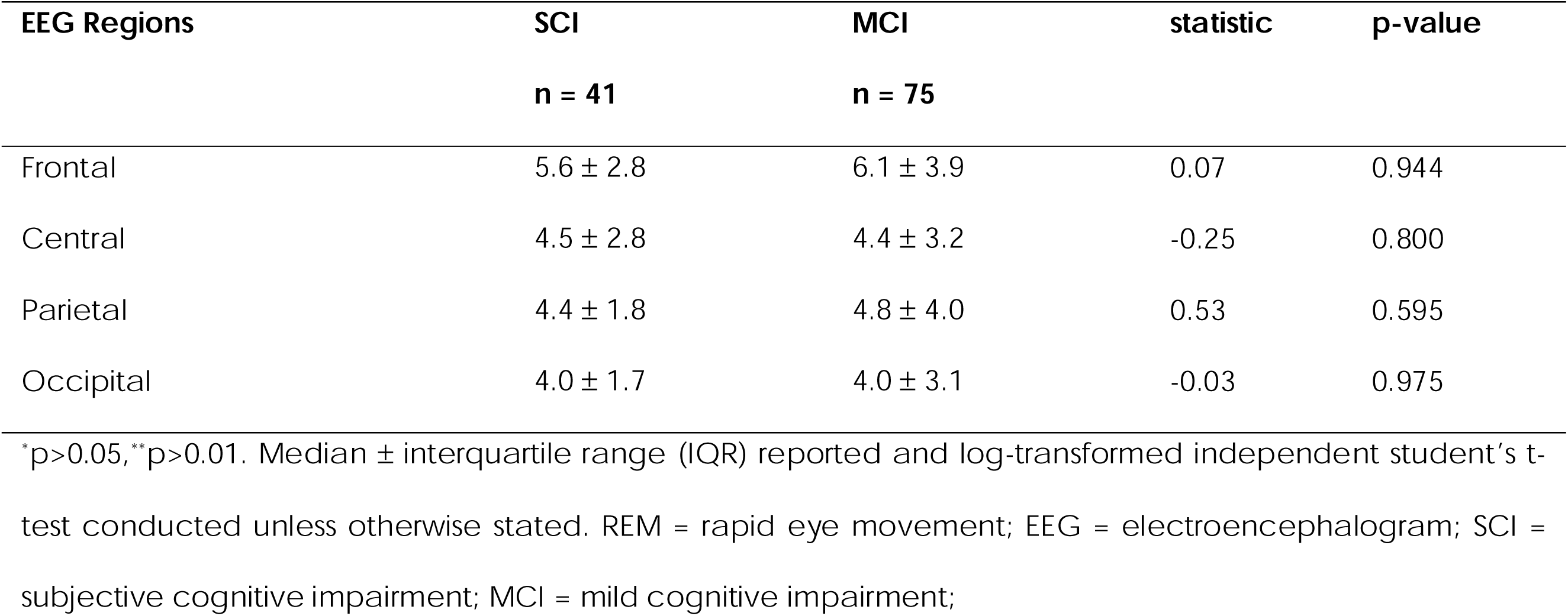
REM EEG slowing differences between SCI and MCI groups.

| EEG Regions | SCI<br>n = 41 | MCI<br>n = 75 | statistic | p-value |
| --- | --- | --- | --- | --- |
| Frontal | 5.6 ± 2.8 | 6.1 ± 3.9 | 0.07 | 0.944 |
| Central | 4.5 ± 2.8 | 4.4 ± 3.2 | -0.25 | 0.800 |
| Parietal | 4.4 ± 1.8 | 4.8 ± 4.0 | 0.53 | 0.595 |
| Occipital | 4.0 ± 1.7 | 4.0 ± 3.1 | -0.03 | 0.975 |

\*p>0.05,\*\*p>0.01. Median ± interquartile range (IQR) reported and log-transformed independent student's t-test conducted unless otherwise stated. REM = rapid eye movement; EEG = electroencephalogram; SCI = subjective cognitive impairment; MCI = mild cognitive impairment;

In the unadjusted model, frontal (β = −0.23, *95% CI* [-0.41, −0.05], *p* = 0.014, *q* = 0.018, *n* = 114), central (β = −0.26, *95% CI* [-0.44, −0.08], *p* = 0.006, *q* = 0.018, *n* = 116) and occipital (β = −0.24, *95% CI* [-0.42, −0.06], *p* = 0.010, *q* = 0.018, *n* = 115) REM EEG slowing predicted total NbM volume after FDR correction. However, the parietal region was not significantly associated with NbM volume (β = −0.17, *95% CI* [-0.39, −0.04], *p* = 0.104, *q* = 0.104, *n* = 88). In adjusted models (controlling for age, sex, OSA severity and MRI sequence), statistically significant associations between REM EEG slowing and NbM were maintained for the frontal (β = −0.25, *95% CI* [-0.43, −0.07], *p* = 0.008, *q* = 0.010, *n* = 114), central (β = −0.27, *95% CI* [−0.45, −0.09], *p* =0.004, *q* = 0.010, *n* = 116) and occipital regions (β = −0.25, *95% CI* [−0.43, −0.07], *p* = 0.008, *q* = 0.010, *n* = 115).

Subsequent analyses examined whether the association between REM EEG slowing and NbM volume differed by cognitive classification. Before correction for multiple comparisons, significant interactions were observed at frontal (*b* = −4.21, *df* = 108, *p* = 0.046) and occipital regions (*b* = −7.01, *df* = 109, *p* = 0.031). However, none of the interaction effects remained statistically significant after FDR correction across the four regions (all FDR-adjusted *p* > 0.05).

We conducted exploratory within-group analyses within the cognitive subgroups which revealed differential findings by group. Within the SCI group, REM EEG slowing and NbM volume were not statistically significant at region (all FDR-adjusted *p* > 0.05). In contrast, greater REM EEG slowing was associated with lower NbM volume in all regions for the MCI group (all FDR-adjusted *p* < 0.05). See Figure 1 for diagnostic group-stratified scatterplots. The effect sizes were moderate in magnitude, with standardised coefficients ranging from β = -0.30 to -0.35. For full b-values, uncorrected and corrected p-values, see Table 4.

**Figure 1.**
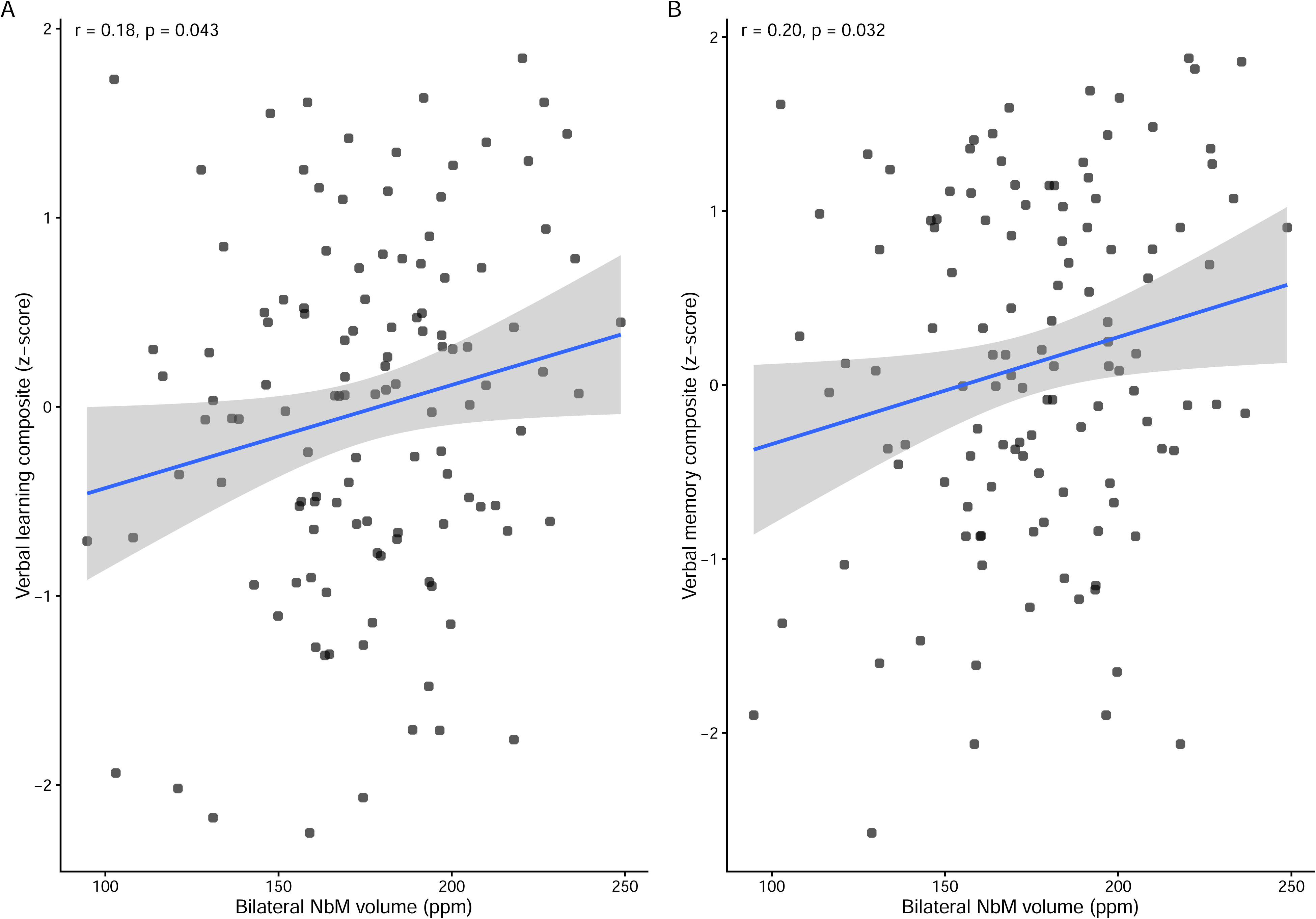
Scatterplots of REM EEG slowing ratio across four scalp regions and Nucleus Basalis of Meynert (NbM) volume with linear regression lines and 95% confidence intervals stratified by diagnostic group. **(A)** Frontal region averaged over available F3, F4, Fz electrodes; **(B)** Central region averaged over available C3, C4, Cz electrodes; (**C)** Parietal region derived from Pz electrode; **(D)** Occipital region averaged over available O1 and O2 electrodes. SCI = subjective cognitive impairment; MCI = mild cognitive impairment; REM = rapid eye movement; EEG = electroencephalogram; NbM = Nucleus basalis of Meynert. TIV = total intracranial volume. Ppm = parts per million.

We next examined if sex or OSA severity moderated the association between REM EEG slowing and NbM volume with resultant analyses showing that they did not moderate the relationship at any electrode, either before or after FDR correction (all *p* > 0.05 and FDR-adjusted *p* > 0.05) (for full models, see Supplementary Table 1 for sex interaction models and Supplementary Table 2 for OSA severity interaction models).

To determine whether the observed associations were specific to the NbM volume, we examined the associations between REM EEG slowing and the bilateral Ch1-3 volumes. It was not associated with REM EEG slowing at any electrode after adjustment for sex, age, OSA and MRI sequence, either before or after FDR correction (all *p* > 0.05; FDR-adjusted *p* > 0.05). See Supplementary Table 3.

To assess whether the observed associations were specific to REM sleep, we examined associations between non-REM EEG slowing and bilateral NbM volume. A greater non-REM EEG slowing ratio at central, occipital and parietal electrodes was linked to lower NbM volume before FDR corrections (*p* < 0.05). However, none of these associations survived FDR corrections (FDR-adjusted *p* > 0.05). For full unadjusted and adjusted models, see Supplementary Table 4.

Our exploratory univariate analyses found that smaller NbM volume was associated with longer sleep latency (*p* < 0.05). There were no other associations between total NbM volume and sleep macroarchitecture (all *p* > 0.05, see Table 5 for the full correlation matrix). Furthermore, in an attempt to replicate previous findings^31^, we examined the relationship between NbM volume and sleep macroarchitecture in a subsample of individuals with aMCI (*n* = 29). Again, there was no significant relationship between NbM volume and sleep macroarchitecture. Lastly, smaller NbM volume was associated with poorer learning (Figure 2A) and memory (Figure 2B) composite scores (*r* = 0.184, *p* = 0.042; *r* = 0.196, *p* = 0.032, respectively), though notably these were small effects.

**Figure 2.**
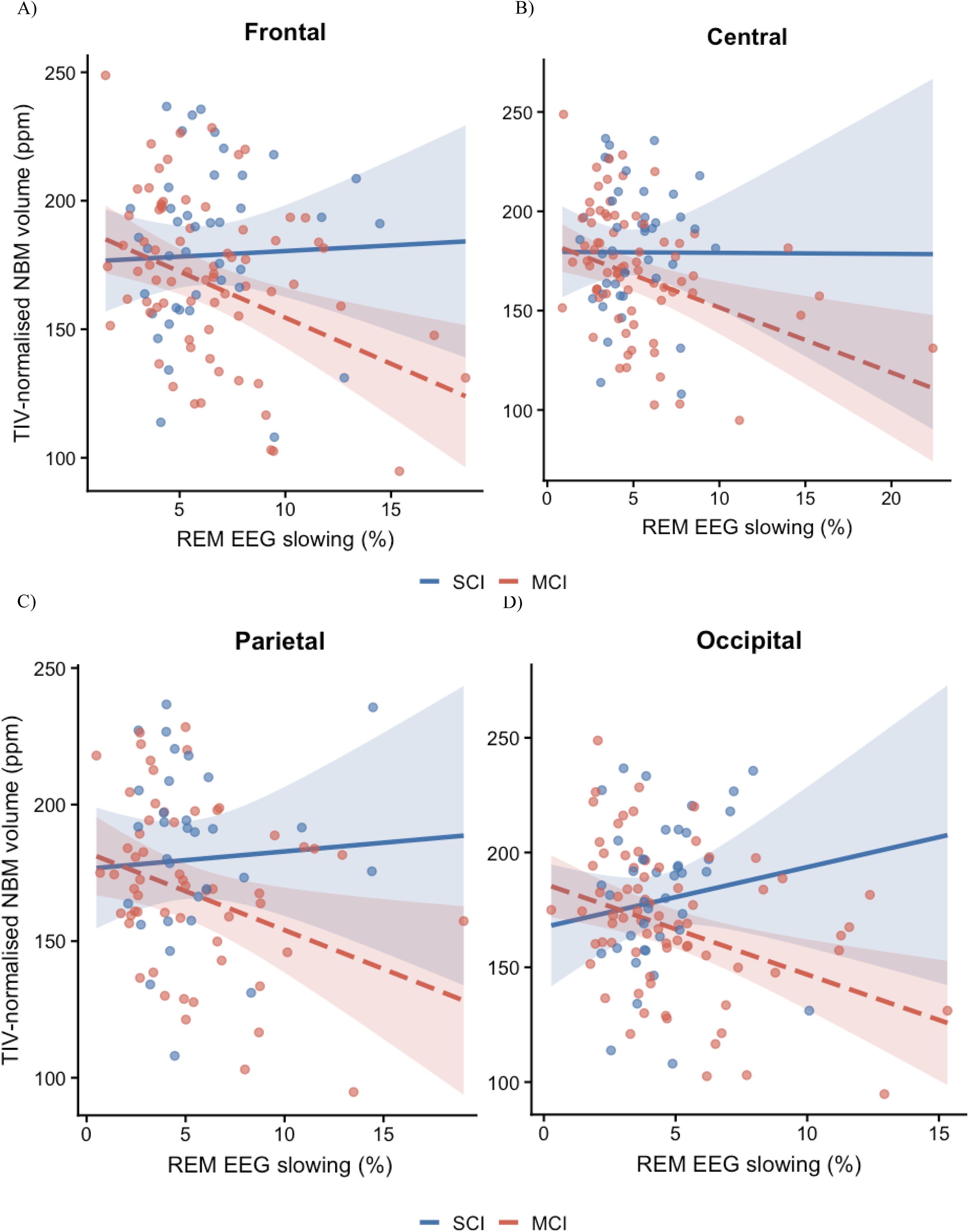
Associations between verbal learning and memory performance and nucleus basalis of Meynert (NbM) volume. Scatterplots show the relationships between NbM volume and **(A)** verbal learning composite *z*-scores and **(B)** verbal memory composite *z*-scores. Solid lines represent linear regression fits, with shaded areas indicating 95% confidence intervals. Pearson’s correlation coefficients were used to assess the strength and direction of the associations.

## Discussion

In the present study, we sought to determine whether REM EEG slowing is associated with NbM volume in older adults with SCI and MCI and whether cognitive status moderated this relationship. Aligned with our primary hypothesis, greater REM EEG slowing was associated with smaller NbM volumes, independent of age, sex, OSA severity and MRI sequence. None of our secondary hypotheses were supported. That is, neither sex or OSA severity moderated the association between REM EEG slowing and NbM volumes. Although we observed an interaction between REM EEG slowing and cognitive status (SCI vs. MCI), this did not survive FDR adjustment. Neither OSA nor sex moderated the association between REM EEG and NbM volume. Exploratory analyses further showed that a smaller NbM volume was associated with longer sleep latency and poorer learning and memory composite scores. Overall, these findings support an association between REM EEG slowing and NbM volume in older adults with SCI and MCI.

The present study provides novel evidence that increased REM EEG slowing is associated with lower NbM volume in clinical samples at-risk and high risk for dementia. This extends prior work linking REM EEG slowing to cholinergic denervation, measured with PET, in a similar cohort of older adults with and without MCI.^10^ Whereas earlier studies utilised molecular imaging, our findings show that structural alterations in the NbM, a major source of acetylcholine,^32^ are also related to altered REM sleep neurophysiology. Importantly, our findings suggest that REM EEG slowing is specific to NbM integrity, as there were no associations between non-REM EEG slowing ratio and the NbM, nor with REM EEG slowing and Ch1-3 volume. The specificity of our findings is consistent with evidence from animal models. In mice, cholinergic basal forebrain neurons fire maximally during REM sleep and are virtually silent during slow-wave sleep.^33^ Their neuronal firing rates are positively associated with fast-frequency activity and negatively associated with delta activity, consistent with a role of NbM cholinergic signalling in maintaining cortical activation during REM sleep. Taken together, REM EEG slowing appears to be a specific marker of cholinergic integrity that can be measured at the scalp level, highlighting its potential relevance as a sleep biomarker of early neurogenerative risk.

Although the interaction with cognitive status did not survive FDR corrections, post-hoc within-group regression models suggested that the effects of REM EEG slowing and NbM is driven by the MCI group. This pattern is aligned with the notion that the MCI stage represents a period of increased vulnerability of the basal forebrain to neurodegeneration.^34^ Neuropathological and neuroimaging evidence indicate that cholinergic neuronal loss and NbM atrophy are more pronounced in individuals with MCI than in cognitively healthy older adults.^35^ Consistent with this literature, our findings suggest that the relationship between REM EEG slowing and NbM volume becomes more apparent in the context of structural degeneration, with the MCI group exhibiting reduced NbM volume relative to the SCI group. This may indicate that REM EEG slowing reflects a functional consequence of structural and molecular alterations in the BF cholinergic system in the early prodromal stages of dementia.^10^ However, due to the cross-sectional nature of the current study, we cannot rule out that REM EEG slowing may emerge concurrently with BF cholinergic changes, and that other underlying disease pathologies (e.g. amyloid, tau) may drive these alterations. We also did not observe a moderating effect of sex or OSA severity, suggesting there may be no sex differences and effect of OSA severity on the observed relationship between REM EEG slowing and NbM volume. Future research should build on these findings and explore the longitudinal relationship between REM EEG slowing and the BF cholinergic system in dementia cohorts.

In both the whole sample and those with amnestic MCI, NbM volume was associated with sleep latency but no other sleep macroarchitecture outcomes, including REM onset latency and REM sleep duration, or OSA. This contrasts with a prior study that reported significant associations between NbM volume and a reduced percentage of REM sleep in participants with amnestic MCI.^31^ One potential difference in our studies is that the prior study excluded individuals with OSA, whereas our cohort included OSA participants. Prior studies have demonstrated that OSA influences REM sleep macroarchitecture,^36^ therefore, the presence of OSA in our cohort may blur the relationship between NbM volume and REM macroarchitecture. Unfortunately, we were underpowered to further exclude OSA in our sub-sample of 29 participants with amnestic MCI. Additionally, we did not observe a link between OSA and NbM volume, a finding that diverges from previous animal studies linking nocturnal hypoxemia and the BF cholinergic system.^37^ The disconnect may stem from the rodent work, which suggested the relationship between OSA and the BF was mediated by inflammatory processes. Therefore, without an accompanying increase in neuroinflammation, degeneration of cholinergic BF neurons may not occur in those with OSA. Unfortunately, we did not include measures of neuroinflammatory markers, and future human studies incorporating markers of neuroinflammation and Alzheimer’s disease biomarkers will be critical for determining whether the associations observed in animal models translate to clinical populations. Nonetheless, our findings extend prior studies by demonstrating that slowing of brain activity during REM sleep may precede or be more sensitive to alterations in NbM volume than the REM-related macroarchitecture changes observed in prior studies, even in the presence of OSA.

The current findings have several research and clinical implications. First, as previously mentioned, REM EEG slowing may serve as a cost-effective and scalable marker of cholinergic structural integrity in older adults at-risk for dementia. Clinically, incorporating REM spectral power measures into sleep assessments could enhance early risk stratification, particularly in populations in which NbM vulnerability may be critical (prodromal Alzheimer’s disease and Lewy Body dementia). It could potentially inform cholinesterase inhibitor therapy, and prior literature has suggested that cholinesterase inhibitor therapy may reduce REM EEG slowing in those with mild-moderate Alzheimer’s disease.^38^ Longitudinal multimodal studies will be particularly informative. Repeated MRI and polysomnography could determine whether progressive NbM volume loss is accompanied by increasing REM EEG slowing and whether either measure predicts subsequent cognitive decline or conversion to dementia. The addition of amyloid, tau, α-synuclein and cholinergic biomarkers would help determine the pathological specificity of the association. Pharmacological studies could also examine whether changes in cholinergic signalling modify REM EEG slowing, although treatment-related normalisation of EEG activity would not necessarily demonstrate reversal of the underlying neurodegenerative process. Finally, future work should evaluate the generalisability of these associations to other clinical groups like dementia with Lewy bodies or Alzheimer’s disease, where NbM pathology and REM sleep disturbances are both prominent.

Several limitations and strengths of the current study are worth noting. Unfortunately, the cross-sectional study design precludes causal or temporal inferences about the relationships among REM EEG slowing, NbM volume and cognitive impairment. In addition, our reliance on structural MRI alone limits insights into functional and molecular mechanisms; integrating multimodal imaging approaches, such as cholinergic PET radiotracers,^39^ resting-state fMRI, diffusion-weighted imaging, as well as amyloid and tau markers, will be critical for fully elucidating the neurobiological pathways linking sleep EEG dynamics, cholinergic integrity and cognition. Nonetheless, our study leverages gold-standard, in-laboratory polysomnography to characterise sleep microarchitecture within a clinically representative cohort. Critically, our study includes older adults with OSA, as our prior work has demonstrated that up to 76% of older adults with SCI and MCI also present with OSA.^12,40^ This strengthens the ecological validity of our findings and supports the applicability of REM EEG slowing to real-world clinical settings, such as memory clinics.

This study provides new evidence that greater REM EEG slowing was associated with lower NbM volume in older adults with SCI and MCI. This finding links altered REM cortical activity with the structural integrity of a major component of the basal forebrain cholinergic system and extends recent molecular imaging evidence relating REM slowing to cortical cholinergic denervation. While further validation is needed, these findings indicate that REM EEG features may offer insight into NbM integrity and broader cholinergic system changes in aging. More broadly, this work highlights REM sleep physiology as a potentially informative window into early neurobiological processes relevant to dementia risk, warranting continued longitudinal and multimodal investigation.

## Data Availability

All data produced in the present study are available upon reasonable request to the authors

## Data availability

The data that support the findings of this study are available from the corresponding author, upon reasonable request.

## Acknowledgements

We extend our sincere gratitude to all the participants who generously contributed their time to this study. We also thank the dedicated administrative and clinical staff at the Healthy Brain Ageing Clinic for their ongoing support, with special thanks to our research managers, Dr Simone Simonetti and Ms Lisa Vizza. We acknowledge the administrative and clinical staff at the Woolcock Institute of Medical Research for their expertise and assistance with data collection throughout this research. Lastly, we acknowledge the use of Grammarly to check for grammatical correctness and sentence structure.

## Supplementary material

Supplementary material is available at *Brain* online.

